# Structural, Social, and Contextual factors influencing COVID-19 vaccine uptake: A qualitative methods study among Healthcare Workers and Older People in Uganda

**DOI:** 10.1101/2023.07.10.23292213

**Authors:** Sande Slivesteri, Agnes Ssali, Ubaldo M Bahemuka, Denis Nsubuga, Moses Muwanga, Chris Nsereko, Edward Ssemwanga, Asaba Robert, Janet Seeley, Alison Elliott, Eugene Ruzagira

**Affiliations:** MRC/UVRI & LSHTM Uganda Research Unit, Wakiso, Uganda; Entebbe Regional Referral Hospital, Wakiso, Uganda; Villa Maria Hospital, Kalungu, Uganda; Our Lady of Consolata, Kisubi Hospital, Wakiso, Uganda; Department of Global Health and Development, London School of Hygiene and Tropical Medicine, London, UK; Department of Clinical Research, London School of Hygiene and Tropical Medicine, London, UK; Department of Infectious Disease Epidemiology, London School of Hygiene and Tropical Medicine, London, UK

**Keywords:** Healthcare Workers, older people, COVID-19 vaccines, barriers, facilitators, uptake.

## Abstract

**Background:** The COVID-19 vaccine rollout program in Uganda was launched in March 2021 with Healthcare Workers (HCWs), older persons (≥50 years), and persons with chronic conditions as priority groups for vaccination. To inform the vaccine rollout efforts, we set out to explore the social and structural factors that influenced the uptake of COVID-19 vaccines among HCWs and older people in Uganda.

**Methods:** Between September and October 2021, we conducted 33 in-depth interviews with 25 HCWs aged 21–63 years from three hospitals from two districts in the central region of Uganda and eight older people from communities in Wakiso district. Selection was purposive based on sex, occupation, education, cadre of HCWs (doctors, nurses, laboratory technologist, hospital support staff, administrator) and vaccination status. We explored participants’ knowledge, beliefs, personal experiences, barriers, and facilitators to vaccine uptake and suggestions for future COVID-19 vaccine rollout. Interviews were audio-recorded, data was transcribed and translated from the local language, coded, and analysed by themes.

**Results:** Twenty-two of the 25 (88%) HCWs and 3 of the 8 (38%) older people had received at least one dose of the COVID-19 vaccine at the time of interview.

The structural facilitating factors to vaccine uptake included access to correct information, fear of a risky work environment, and mandatory vaccination requirements especially for frontline HCWs. Old age, chronic health conditions, and the fear of death are contextual facilitating factors, while influence from leaders was the main social facilitating factor.

Myths and misconceptions about COVID-19 vaccines and the fear of side effects were common social barriers to vaccine uptake among HCWs and older people. Long distances to vaccination centres, vaccine stock-outs, and long queues at the vaccination centres were specific barriers to vaccine uptake for older people. The prerequisite of signing a consent form was a specific structural barrier for the HCWs. Transport challenges linked to long distances to the vaccination centres, for older people, and having underlying chronic health conditions, for both older people and HCWs, were the reported contextual factors.

**Conclusion:** Future roll out of new vaccines should have a comprehensive information dissemination strategy about the vaccines. Improved access to vaccines through community outreaches, reliable vaccine supply and addressing vaccine misinformation, may enhance COVID-19 vaccine uptake in Uganda and other future mass vaccination campaigns.

## Background

By June 2023, the coronavirus (COVID-19) pandemic had caused 12.2 million confirmed cases (256,542 deaths) in Africa and 170,544 confirmed cases (3,632 deaths) in Uganda [1]. The pandemic placed a considerable strain on the public health system particularly in the early phase in 2020. Besides lockdown prevention measures causing great mobility challenges in accessing health facilities, many public health facilities scaled down on antenatal, HIV, vaccination, and other health services to focus on COVID-19 especially in 2020-2021 [2-4].

Several vaccines have proved to be safe and effective against laboratory-confirmed SARS-CoV-2 virus infection and symptomatic or severe COVID-19 disease [5-7]. Since December 2020, several countries initiated COVID-19 vaccine rollout programs, with 67% of the world population fully vaccinated and 31.5% vaccinated with at least one booster or additional dose by June 2023. Vaccination rollout has been reportedly slower in Africa, with only 49.7% of the African population fully vaccinated [1, 8]

In Uganda, COVID-19 vaccine rollout was launched on 10^th^ March 2021 targeting at-risk groups: HCWs, security personnel, people aged 50 years and above, and those aged 18–50 years with comorbidities [9-11]. COVID-19 vaccination was scaled up to target all persons aged 18 years and above and children of 12 years above in 2022 [11, 12].

Findings from a study in South Africa indicated that acceptance of a new COVID-19 vaccine was influenced by different factors including age, employment status, urbanity, and geographical location [13]. Another study among high-risk populations in Uganda showed that 70% of the participants were open to receiving the COVID-19 vaccine with the probabilities being four times higher in men versus women [9].

However, other studies reported concerns about the vaccine’s expedited development and approval process, potential side effects, and efficacy [14-17]. Such concerns contribute to vaccine hesitancy, which has been reported as a significant challenge to disease prevention and control worldwide. The WHO describes vaccine hesitancy as the delay in the acceptance or complete refusal of vaccines [18]

Structural, social, and contextual factors may contribute to vaccine confidence and affect the uptake of the COVID-19 vaccine among HCWs and older people in Uganda. We set out to explore the structural, social, and contextual factors that influence the uptake of COVID-19 vaccines among HCWs and older people aged ≥ 50 years using the Social Ecological Model (SEM) to structure our analysis.

### Theoretical framework

The Social Ecological Model (SEM) provides a framework for exploring the relationship between social and structural factors and the physical environment and how such factors influence and shape the decision-making process of individuals, including their health-seeking behaviours [19, 20]. The model been used to describe individuals’ behaviours using measurements comprising of intrapersonal (within the individual), interpersonal (between individuals), institutional, community and public policy factors to provide a framework for understanding the interplay between these levels and how they shape or influence an individual’s actions and choices[20, 21].

Using this framework, we explored participants’ knowledge, beliefs, and attitudes about COVID-19, personal experience about COVID-19 vaccines, facilitators and barriers to vaccine uptake, and opinions on the future of vaccine rollout. See figure 1.

**Figure 1:**
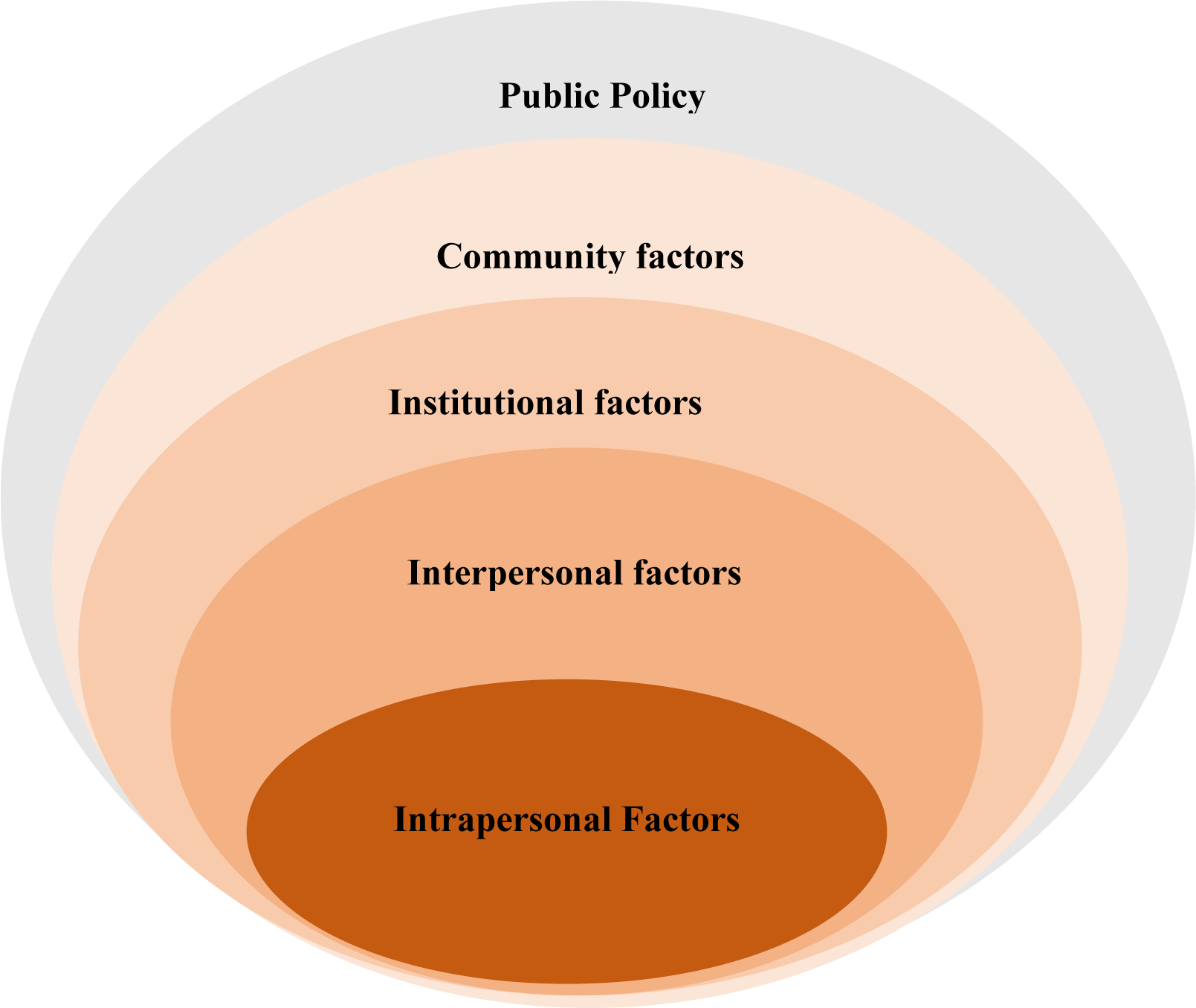
The social ecology of health promotion interventions. Health Education Quarterly, 15(4):351-377. Adapted from McLeroy, K. R., Steckler, A. and Bibeau, D. (Eds.) (1988).

The intrapersonal level includes individual characteristics, such as age, education, income, and health history, Interpersonal level comprise of relationship and interaction with others in a person’s closest social circle, such as friends, partners, and family members, all of whom influence a person’s behaviours including health-seeking behaviours. The institutional layer that includes the social organisation characteristics, rules, and regulations for operation. The community layer explores the settings in which people have social relationships, such as schools, workplaces, and neighbourhoods, and seeks to identify the characteristics of these settings that affect health. Finally, the public policy layer explains the broad societal factors that favour or impair health including state regulations.

## Methods

### Study design and setting

This was a qualitative methods study conducted among a subset of participants enrolled between May and October 2021 in a prospective study to explore acceptability and immunogenicity of COVID-19 vaccines among 597 HCWs and 150 older people (≥50 years) in Uganda. The HCWs were enrolled from three hospitals: I) Entebbe regional referral public hospital, located in Entebbe municipality, 44km from Kampala, the capital city of Uganda; II) Our Lady of Consolata Kisubi Hospital, a Private Not-For-Profit (PNFP) hospital located approximately 28km from Kampala, and Villa Maria Hospital, a PNFP hospital in Kalungu district, approximately 138km from Kampala. The older people were participants in the Well-being of Older People Study (WOPs) that has been ongoing in the semi-urban Wakiso district and the rural Kalungu district for 12 years. The objective of WOPs is to describe the roles, health problems (physical and mental) and social well-being of older people directly and indirectly affected by HIV/AIDS, with special attention to the effects of the introduction of Antiretroviral Therapy (ART). In the current study, we enrolled HCWs from the three hospitals and older people from Wakiso.

### Study participants, sampling, and data collection

Using the participant identification logs of COVID-19 vaccine acceptability and immunogenicity study, we purposively identified and invited 33 individuals who had been enrolled in the preceding three weeks to participate in the in-depth interviews. Selection was based on sex, occupation, education, cadre of the HCWs (Doctors, Nurses, Laboratory technicians, hospital support staff and administrators) and vaccination status. Those selected included 25 HCWs (Kisubi hospital-8; Entebbe regional referral hospital-8; Villa Maria hospital-9) and 8 WOPs.

Social science research assistants (RAs) contacted the selected individuals via phone calls and requested their participation in the in-depth interviews. The RAs and the identified participants agreed on the date, time, and convenient venue to discuss study information, and if they consented, interviews were conducted on the same day.

Interviews were conducted using a semi-structured interview guide that included the following topics: knowledge, beliefs, and attitudes towards COVID-19 vaccines; enabling factors and barriers to uptake of COVID-19 vaccines; and personal views on the future of the COVID-19 vaccine rollout programme. The interviews were audio recorded if the participant agreed, or notes were taken if the participant declined voice recording. Interviews were conducted in the local vernacular language (Luganda). A few interviews were conducted in English for the participants who preferred this. Each interview lasted about 45 minutes. A senior social scientist conducted regular debriefing meetings with the RAs to review the completeness of the data and identify areas to improve on in subsequent interviews.

### Data management and analysis

Anonymized audio files were transferred onto encrypted password-protected computers, transcriptions and translations were done by the RAs, and anonymized transcripts were transferred to a secure data server at the Medical Research Council /Uganda Virus Research Institute and London School of Hygiene and Tropical Medicine (MRC/UVRI and LSHTM) Uganda Research Unit. Data analysis was iterative with the research team sharing and discussing transcripts to identify and agree on common codes. A codebook was developed describing the meaning of each code. All the transcripts were coded using both inductive and deductive coding and patterns from the data led to the themes identified in this paper informed by the layers from the SEM.

### Ethical considerations

The study was approved by the Uganda Virus Research Institute Research Ethics Committee (UVRI REC GC/127/21/03/813), the Uganda National Council for Science and Technology (UNCST SS767ES), 27-04-2021, and the London School of Hygiene and Tropical Medicine Research Ethics Committee (25997). We obtained administrative clearance from all the collaborating hospitals to conduct the study. Written informed consent to participate in the interviews was obtained at enrolment time in the main study of acceptability and immunogenicity of COVID-19 vaccines. Before each interview, RAs verbally checked to confirm that participants were still interested in taking part in the in-depth interview. All interviews were conducted in a safe and private place to ensure participants’ privacy and confidentiality.

## Results

### Demographic characteristics of study participants

Among the eight older people, the mean age was 68.8 (SD 6.7) years, 50% were female, 75% were Christian, 50% had attained at least secondary education level and 37.5% had received at least one dose of the vaccine (Table 1). Among the 25 HCWs, the mean age was 37.8 (SD 11.2) years, 32% were female, all were Christian, 68% had at least Diploma/bachelor’s degree or other higher-level education, and 92% had received at least one dose of SARS-CoV2 vaccine (Table 1).

**Table 1:** Characteristics of study participants

|  | **Older persons**<br>**(N=08)** | **Healthcare workers**<br>**(N=25)** |
| --- | --- | --- |
| **Characteristics** | **n (%)** | **n (%)** |
| **Sex** |  |  |
| Female | 4 (50) | 8 (32) |
| Male | 4 (50) | 17 (68) |
| **Mean age (SD)** | 67.8 (6.7) years | 37.8 (11.02) years |
| **Age group (years)** |  |  |
| 20-30 | - | 8 (32) |
| 31-40 | - | 8 (32) |
| 41-50 | - | 6 (24) |
| 51-60 | 1 (12.5) | 2 (8) |
| 61-70 | 4 (50) | 1 (4) |
| 71-80 | 3 (37.5) | - |
| **Religion** |  |  |
| Christian | 6 (75) | 25 (100) |
| Muslim | 2 (25) | - |
| **Highest Education Levels** |  |  |
| Incomplete primary | 2 (25) | - |
| Complete primary | 2 (25) | - |
| Incomplete O' level | - | 1 (4) |
| Complete O' level | 2 (25) | 4 (16) |
| Post O' level certificate and Vocational | 2 (25) | 2 (08) |
| Diploma | - | 4 (16) |
| Degree and above | - | 14 (56) |
| **Occupation** |  |  |
| Market vendors | 2 (25) |  |
| Small business owners | 2 (25) |  |
| Farmer | 1 (12.5) |  |
| Teacher | 1 (12.5) |  |
| Mechanic | 1 (12.5) |  |
| Volunteer counsellor | 1 (12.5) |  |
| Doctors | - | 3 (12) |
| Nurses | - | 6 (24) |
| Laboratory technicians | - | 3 (12) |
| Hospital Administrators | - | 2 (16) |
| Human resources | - | 2 (8) |
| Physiotherapist | - | 1 (4) |
| Radiographer | - | 1 (4) |
| Health economist | - | 1 (4) |
| Security guards | - | 3 (12) |
| Cleaners | - | 2 (8) |
| Records manager | - | 1 (4) |
| **District (setting)** |  |  |
| Kalungu (rural) | - | 9 (36) |
| Wakiso (semi-urban/urban) | 8 (100) | 16 (64) |
| **Hospitals** |  |  |
| Villa Maria Hospital | - | 9 (36) |
| Entebbe Regional Referral Hospital | - | 8 (32) |
| Our Lady of Consolata, Kisubi Hospital | - | 8 (32) |
| **Vaccination status** |  |  |
| Vaccinated (at least one dose) | 3 (37.5) | 23 (92) |
| Not vaccinated | 5 (62.5) | 2 (8) |

N=number; SD=standard deviation

We present our results guided by the structure of the SEM, which defines human behaviours, interactions across societal levels, and actions. See figure 2.

**Figure 2:**
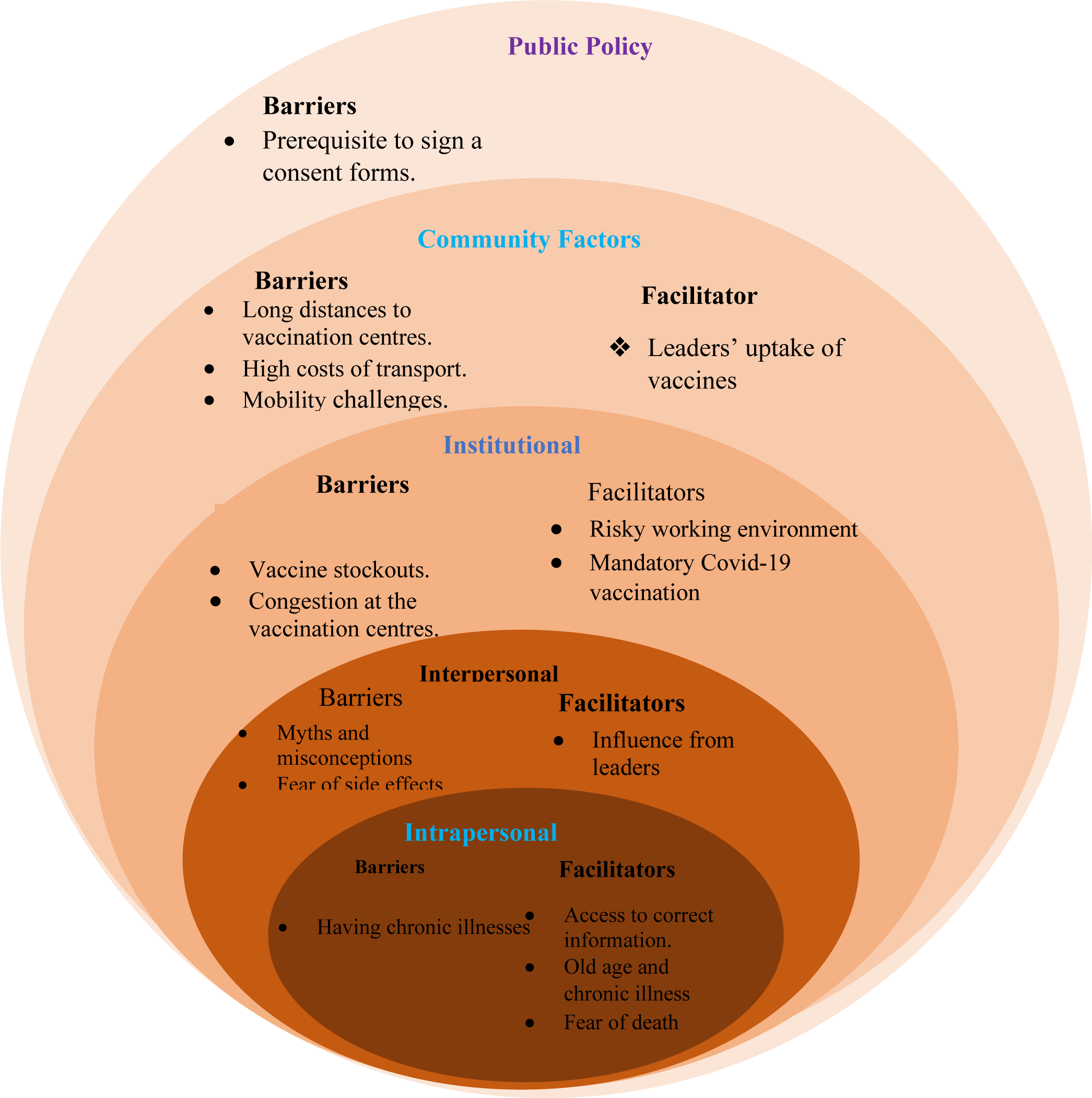
Structural, Social and Contextual factors influencing COVID-19 vaccine uptake.

## Barriers to vaccination

### Intrapersonal barriers

Intrapersonal/individual factors, as described by the SEM as the first level, comprises of knowledge, awareness, attitudes, beliefs, and perceptions. We captured participants’ barriers to vaccine uptake based on their individual attributes, knowledge, beliefs, and attitudes towards COVID-19 vaccines.

Having chronic illnesses such as HIV, diabetes, hypertension, and allergies was described as a barrier to vaccine uptake by HCWs and older people. Participants shared the belief that when a person with such chronic conditions uptake the COVID-19 vaccine, it worsens their already existing chronic health condition, even when people with chronic health conditions are prioritised for the COVID-19 vaccination.

> *"I have a disease called allergy, that is the only reason, why I have not been vaccinated yet"*. HCW, not vaccinated.

The older people reported that HCWs advised people with chronic diseases and conditions to delay receiving the vaccine.

> *“Whenever I get there (to the vaccination centres), the health workers tell me that the blood pressure is high and urge that I should wait for it to reduce before I take the vaccine"*. Older person not vaccinated.

### Interpersonal barriers

Interpersonal/relationship factors are the social norms that exist among individuals, groups, or organizations that can limit or enhance health behaviours. Individuals’ interactions with people close to them or people in their families/communities shape their behaviours, attitudes, and preferences, and influence their decision-making including health-seeking behaviours. The barriers included myths and misconceptions and fear of the vaccine effect.

The myths and misconceptions attributed to social media posts and people’s narrations within homes and communities were reported to have caused fears and worries among the HCWs and older people about the vaccine. The myths and misconceptions that vaccines were intended to kill Africans, cause infertility and linking the COVID-19 vaccines to acts of “devil/satanic worship” were barriers to vaccine uptake. There was a reported misconception that people who received the vaccine would die after two years of being vaccinated. There were also concerns about infertility:

> *“I am not vaccinated because I have had that belief, that COVID-19 vaccines cause infertility”* HCW, not vaccinated.

> *"They would say that in men it (vaccines) causes impotence and in female, you would be barren because it was a new drug, people did not know about it. So, those who had not yet had children said let me first wait and see"* HCW, vaccinated.

> *" I don’t want to be vaccinated because I heard about the infertility thing and the side effects like general body weakness and things of that kind”* HCW, not vaccinated.

The HCWs and older people described their fears of known and anticipated vaccine side effects as barriers to vaccine uptake. They attributed these vaccine fears to several social media posts and community interactions with vaccinated people.

> *“I usually see these videos and children shows them to me like my daughter they have phones, and they show them to me that have you seen. They have said this and that and the vaccine they have brought this time round is fake”* older person, vaccinated.

> *“Because I knew that I was going to get the disease itself introduced in my body, I had some fears that I could develop the disease itself, and you know, knowing that you we got the vaccine without testing whether we COVID-19 or not”* HCW, vaccinated.

HCWs described the out of context interpretation of vaccine side effects by some people impacting the vaccine uptake.

> *"To the patients, when they come, and they do not have a fever, they happen to start feeling fever and severe headache after vaccination. They conclude that they have been given poison”* HCW, vaccinated.

### Institutional Barriers

The SEM encompasses institutional factors, such as organizational formalities and informal characteristics, including operational rules and regulations. Vaccine stockouts and long wating hours are reported to have negatively impacted on the vaccine uptake.

HCWs and older people reported vaccine stock-outs at the vaccination centres as a barrier to vaccine uptake. HCWs reported sending away people who came to the health facilities for vaccination because of vaccine stock-outs.

> *“I went to Kisubi hospital to be vaccinated and there were no vaccines. They gave me a telephone number to call them and check if they have the vaccines. I have called them twice, and the vaccines are not yet available"* Older person, not vaccinated.

Failure to find vaccines at the vaccination centres after travelling long distances was a reported setback to vaccine uptake.

> *"Limited vaccine doses like now people intending to receive the vaccines come, and you tell them that the vaccines are not available, and people move several times to the hospital and do not find the vaccine"*. HCW, vaccinated.

The congestion at vaccination centers, coupled with the lack of special arrangements to facilitate easy access to vaccines for older people, created barriers to vaccine uptake. The older people reported that they could not stand in long queues at the vaccination centres to receive the vaccines.

> *"In the beginning, being a new thing (COVID-19 vaccine), I know that it attracts many people. In my life, I do not want to struggle and be in a long queue like many people standing in the same place. I would not go there (hospital) because people were many people at the vaccination sites"* Older person, not vaccinated.

### Community barriers

With in the SEM, community factors include the interactions and relationships between institutions and individuals within the community. Long distances to vaccination centre coupled with high transport costs and mobility challenges are community barriers reported.

The older people reported transport challenges getting to the vaccination centres within the communities as a barrier to vaccine uptake. The long distances to the vaccination centres and the COVID-19-related public transportation restrictions e.g., the requirement for public transport vehicles to operate at half capacity led to increased transport costs.

> *“I do not have transport to UVRI Entebbe or Kisubi hospital, that is the main reason why I am not vaccinated, nothing else”* Older person, not vaccinated.

Participants emphasised that transport costs coupled with age-related mobility challenges were a major hindrance to vaccine uptake.

> *"The older person has difficulties taking her/him from where she/he is to the vaccination centres, they cannot move, many do not have money for transport,"*. Older person not vaccinated.

Long distances to vaccination centers were reported as a challenge by HCWs and older people. They highlighted that COVID-19 vaccines were not available in lower-level health facilities, such as Health Centres II, which are in closer proximity to the population. HCWs emphasized that vaccines were only accessible at Health Centre III and hospital-level health facilities. This was reported to have influenced vaccine access because people had to move long distances to the vaccination centres.

> *"The distance to the vaccination centres is long."* Older person not vaccinated.

> *"Some people’s homes are too far from the health centres that are giving the vaccines because there are villages that are far from the health facilities and vaccines are not available in those communities”*. HCW, Vaccinated.

### Public policy barrier

At the final level, the SEM explains how state laws and policies provides a general framework shaping behaviours including health seeking decisions.

Some HCWs complained about the unusual process of accessing the vaccine that requires one to sign a consent form. This requirement caused fears and doubts about the safety of the COVID-19 vaccine among those intending to get vaccinated because it was unusual.

> *“Why do they first want us to consent? So, they mean that if I have any problem, there is nowhere I can report because I consented. So, most people fear that consent form*. HCW, vaccinated.

## Facilitators to vaccination

### Intrapersonal facilitators

HCWs and older people reported having access to correct information on COVID-19 vaccines as a facilitator to vaccine uptake. The World Health Organization (WHO) COVID-19 updates, the Continuous Medical Education series (CMEs), the updates from the Ministry of Health, training by the District Health Teams (DHT), the internet, and social media were reported as specific sources of information for HCWs. The news media (Television and radio) were the information sources for older people and some HCWs. Some HCWs reported being approached by the hospital management, convincing them to uptake the vaccine.

> *“Management had to sit down and singled us out (unvaccinated), and they actually brought one of the persons who work with WHO mainly on vaccination, to talk to us, to remove the doubts we had”*. HCW, vaccinated.

For some participants, being of old age and having chronic health conditions was described as one of the factors that prompted them to uptake the vaccine. Participants with diabetes, persons living with HIV, and hypertension reported having accepted to receive the vaccine because of fear of the detrimental effects of COVID-19 in unvaccinated persons with these conditions.

> *"My age prompted me and the underlying diseases I have. First, I have diabetes, I have high blood pressure, and I am on antiretroviral therapy"*, Older person, vaccinated.

> *“I am not very young. Because I realized that if the disease was to strike me, maybe, I would not have a chance to fight it much”*. HCW, Vaccinated.

The belief in the vaccine’s potential to provide protection against the coronavirus played a significant role in motivating HCWs and older individuals to actively seek vaccination.

> *"Fear of death because the moment you see people struggling in the ICU, you will run and take these vaccines”*. HCW, vaccinated.

### Interpersonal facilitators

Influence from leaders who took the vaccine was the leading social factor mentioned by some participants. Participants reported that they gained confidence that the COVID-19 vaccines were safe after witnessing religious, cultural, and political leaders, employers (hospital administrators) and peers receive the vaccine.

> *" As you see the leadership going for the same, (vaccination), you get motivated, like when for example, the minister of health took the jab"*. HCW, vaccinated.

Some hospital administrators reported taking the vaccine to encourage their peers and the people they led to receive it. Other HCWs reported that they went in for vaccination after interacting with their peers who had taken the vaccine and looked normal.

> *"There was no way that I could convince my people (juniors) to go for vaccination, yet I had not done the same"*. HCW, vaccinated.

### Institutional factors

The risky working environment at the hospitals was a facilitator of vaccine uptake for the HCWs. Some HCWs revealed how reluctant they were to receive the vaccine until they were assigned to work directly in COVID-19 patients’ wards. In addition, HCWs reported constant exposure to COVID-19 patients at the hospitals as motivator for getting the vaccine.

> *“I am always exposed. I am always among patients, so I could not think that I could never get the disease. So, I could not take it for granted, because we are always exposed among these patients”*. HCW, vaccinated.

> *"It is very funny. At first, I was a bit hesitant to take the vaccine. There came a time when hospital administration was looking for a staff who are going to perform COVID-19 tests, and I was among the staff selected to perform COVID-19 testing. I started thinking of my safety”*. HCW, vaccinated.

The implementation of mandatory COVID-19 vaccination requirements for HCWs served as a decisive factor for some HCWs, leading them to opt for vaccination to maintain their employment. Some older people reported taking the vaccine in anticipation of future mandatory movement permits in the form of COVID-19 vaccination cards.

> *"So, when management sat, they decided to change the policy, that please, if you are not vaccinated, go home and get vaccinated or if you fall sick, you pay your own bills. It was hard at first until one of the persons; one staff got sick who was not vaccinated”*. HCW, vaccinated.

> *"At first, it was optional, we did not put much stress on everyone getting vaccinated, but as time went on, we made it mandatory"*. HCW, vaccinated.

> *“Even us, the health care workers, currently vaccination is a policy for the institution"*. HCW, vaccinated.

### Participants’ recommendations to reduce barriers to COVID-19 vaccine uptake

Participants’ recommendations to reduce barriers to COVID-19 vaccine uptake included using various communication channels e.g., radio, social media, posters, and megaphones to share information on COVID-19 vaccines (e.g., benefits, safety, and development processes) widely in all the communities, ensuring vaccine availability, conducting targeted vaccination outreaches, putting in place mandatory vaccine policies, initiating in-country (local) manufacturing of vaccines, and financial allowances to HCWs involved in COVID-19 vaccination campaigns.

> *“They should continue educating people and creating awareness. There is no other approach apart from that because if a person understands, he/she will know what exactly is taking place”* Older person, not vaccinated.

> *“The information should continue running on air, in communities, health workers should start going to the communities and do health education in the communities. It should be regular”*. HCW, vaccinated.

Some HCWs underscored the relevance of providing enough information for people to appreciate the benefits of receiving the vaccine and cautioned that mandatory vaccination would instead force people to get fake vaccination certificates without necessarily receiving the vaccine.

> *"The best thing is to counsel somebody to know why he is getting the vaccine. Yes, that is the best way because the policy will be there, and somebody will go to Nasser Road (a street in Kampala that is notorious for being a source of forged documents) and get a fake card”*. HCW, vaccinated.

The HCWs and older people recommended regular and consistent vaccine supply to the vaccination centres.

> *"They have to increase the quantity of the vaccines. At least this time, people want the vaccines, but they are not available. So, they should make them available"*. HCW, vaccinated.

Targeted vaccination outreaches that take the vaccines nearer to the people were recommended as a facilitator of vaccination uptake.

> *“The government should bring the vaccines and try to bring it close to people in the communities”*. Older person, vaccinated.

> *"They should put vaccine outreaches such that people can reach every person far deep down on the ground as they have done for HIV"*. HCW, vaccinated.

The HCWs and older people recommended local vaccine manufacture as opposed to vaccine importation.

> *"Uganda should work on manufacturing [its] own vaccine. [Make] Our own vaccine that is manufactured in Uganda, not from outside, as people fear vaccines from outside, which they say [are feared because] the whites want to kill them"*. HCW, vaccinated.

The recommendation for local vaccine manufacture is believed to enhance vaccine availability, supply reliability, and foster confidence in vaccine safety.

## Discussion

Our study highlights several individual, interpersonal, institutional, community, and public policy level barriers and facilitators of COVID-19 vaccine uptake among HCWs and older persons in the early phase of vaccination roll-out in Uganda.

Among the barriers, at an intrapersonal level, we found out that having chronic illnesses was a barrier and at the same time a facilitator to vaccine uptake, while fear of vaccine side effects, myths and misconceptions were reported as interpersonal barriers to vaccine uptake. At the institutional level, we found out that vaccine stockout and congestion at the vaccination centres were major health facility operational barriers. The high transports costs, long distance to the vaccination centres and mobility challenges were community barriers while the prerequisite of signing consent forms was a public policy related barrier to vaccine uptake.

Access to correct information and old age were reported as positive intrapersonal facilitators while fear of side effects, and the influence of leaders were reported to be community facilitators to vaccine uptake. The risky working environment (hospitals) and the hospital vaccination requirements were reported as institutional requirements that promoted vaccine update.

Myths, concerns, and conspiracy theories about COVID-19 vaccines e.g., causing infertility, impotence, and weakening the body’s immunity were major interpersonal barriers to vaccine uptake. Myths and conspiracy theories are well-recognized drivers of vaccine hesitancy globally [22-25]. As exposure to reliable information is a crucial intrapersonal tool for assisting people in making informed judgments [26-28], efforts to actively identify and address COVID-19 misinformation are needed to increase the acceptance of COVID-19 vaccines and other future mass vaccination campaigns, particularly those involving new vaccines.

Old age, presence of chronic health conditions, and working in a hospital setting were facilitating factors for uptake of COVID-19 vaccines in the current study. These findings are consistent with those of other studies [29-31] and are expected since these populations were prioritised for COVID-19 vaccination due to their increased risk of getting infected with SARS-CoV-2 and poor COVID-19 outcomes [32-34]. However, we also found that presence of underlying chronic conditions was the reason for deferral or non-uptake of COVID-19 vaccination for some participants because of fears that vaccination could possibly make these conditions worse. This underscores the need for strategies to increase comprehensive knowledge about the effectiveness and safety of COVID-19 vaccines in the most-at-risk populations.

At the community level, leading by example through the public vaccination of high-profile community leaders or by HCWs getting vaccinated before vaccinating other people facilitated uptake of vaccination. The ecological systems theory emphasizes that human behaviour can be influenced by people who include immediate family members, friends and social networks [35]. Agha et al., 2021 in their research showed that individuals can easily accept or refuse health campaigns depending on the attitude and perception of influential persons in their communities [36].

At the structural level, our study identified several barriers to COVID-19 vaccination uptake that particularly impacted older persons (non-HCWs). These included long distances to vaccination centres and associated transport costs, crowding at the health facilities, and limited vaccine supplies and/stockouts. Similar barriers were reported in other African settings during the early phase of the epidemic [37]. These may have been attributed, at least in part, to limited vaccine supplies, funding shortfalls, a lack of vaccinators, sub-optimal training, inadequate planning, and COVID-19 related disruptions to essential health services [38].

Our results show both support for and caution against mandatory COVID-19 vaccination. Whereas some individuals may be encouraged to get vaccinated to stay in employment, be able to access certain public spaces and/or travel, this approach may have limitations. For example, persons who are strongly opposed to or who are not convinced of the vaccination benefits might opt to use fake COVID-19 certification. Mandatory COVID-19 vaccinations could also have ethical implications. For example, the risk of exacerbating trust breakdowns between employees and their institutions [39].

In the early phase of COVID-19 vaccine roll-out in Uganda, individuals intending to receive the Covid-19 vaccines were required to sign a consent form as a prerequisite [40]. Our results show that this requirement may have negatively impacted uptake of COVID-19 vaccination. Formal consent is not a requirement for other vaccinations in the country. COVID-19 vaccines were first made available under emergency use authorization i.e., a regulatory mechanism to facilitate the availability of unapproved product medical products, including drugs and vaccines, during a public health emergency. The unusual requirement to consent for vaccination coupled with the misinformation about COVID-19 vaccines discussed above, may have heightened concerns about the safety of the vaccines. This finding is in agreement with the study of Bîrsanu., et al that indicated that the unusual informed consent requirement instead caused reduced confidence in the vaccine [41]

### Strengths and limitations of the study

A strength of this study is that we enrolled individuals prioritised for COVID-19 vaccination during the early phase of COVID-19 vaccination campaign and during the second wave of COVID-19 in Uganda (May to October 2021). Thus, study participants’ responses were informed by real-life experiences of the vaccination campaign including receipt of information about, seeking, and receiving the vaccines. A possible limitation is that older participants were all enrolled from Wakiso district, a mostly semi-urban/urban setting. Hence our results may not reflect the lived experiences of rural older persons in relation to COVID-19 vaccine uptake.

## Conclusion

To maximize the success of mass vaccination campaigns, particularly for new vaccines, it is crucial to implement a comprehensive information dissemination strategy that effectively communicates important details about the vaccines being rolled out. By ensuring improved access to vaccines through community outreaches, maintaining adequate vaccine stocks, and providing robust health education to address vaccine misinformation and myths, it is possible to enhance COVID-19 vaccine uptake, particularly in countries where uptake remains low. Moreover, implementing these strategies can potentially increase the uptake of future vaccines, including those designed to combat specific diseases such as Ebola virus in Uganda and similar contexts.

## Data Availability

Because of the characteristics of the data and the consent given by participants, with a
guarantee of anonymity, we are unable to include all the original data. This precaution
is necessary as certain details could potentially compromise the anonymity of the
participants. Hence, the data forming the basis of this paper have been incorporated
into the Supplementary information, ensuring that any mention of names, age, and
specific job identifiers has been excluded.

## Acknowledgments

We would like to express our appreciation to the staff of Villa Maria Hospital, Our Lady of Consolata, Kisubi Hospital, and Entebbe Regional Referral Hospital who participated in this study as well as the entire study team at MRC/UVRI & LSHTM Uganda Research Unit. Special thanks to Dr. Simon Peter Kibira, Senior Lecturer at the College of Health Sciences, School of Public Health, Makerere University, Uganda and Lazaaro Mujumbusi, a Social Scientist at the Medical Research Council/Uganda Virus Research Institute and London School of Hygiene & Tropical Medicine Uganda Research Unit for the invaluable motivation and capacity building that greatly contributed to the inspirations this publication.

## Funding

This work was funded through the Makerere University-Uganda Virus Research Institute Centre of Excellence for Infection and Immunity Research and Training (MUII). MUII was supported through the DELTAS Africa Initiative (Grant no. DEL-15-004). The DELTAS Africa Initiative was an independent funding scheme of the African Academy of Sciences (AAS), Alliance for Accelerating Excellence in Science in Africa (AESA) and supported by the New Partnership for Africa’s Development Planning and Coordinating Agency (NEPAD Agency) with funding from the Wellcome Trust and the UK Government. The work was conducted at the MRC/UVRI and LSHTM Uganda Research Unit which is jointly funded by the UK Medical Research Council (MRC) part of UK Research and Innovation (UKRI) and the UK Foreign, Commonwealth and Development Office (FCDO) under the MRC/FCDO Concordat agreement and is also part of the EDCTP2 programme supported by the European Union. Alison Elliott is supported in part by the NIHR (NIHR134531) using UK aid from the UK Government to support global health research. The views expressed in this publication are those of the author(s) and not necessarily those of the funders or the UK government.

## Notes

### Competing Interest Statement

The authors have declared no competing interest.

### Author Declarations

The study was approved by the Uganda Virus Research Institute Research Ethics Committee (UVRI REC GC/127/21/03/813), the Uganda National Council for Science and Technology (UNCST SS767ES), 27-04-2021, and the London School of Hygiene and Tropical Medicine Research Ethics Committee (25997). We obtained administrative clearance from all the collaborating hospitals to conduct the study. Written informed consent to participate in the interviews was obtained at enrolment time in the main study of acceptability and immunogenicity of COVID-19 vaccines. Before each interview, Research Assistants verbally checked to confirm that participants were still interested in taking part in the in-depth interview. All interviews were conducted in a safe and private place to ensure participants privacy and confidentiality.

